# Booster vaccination to curtail COVID-19 resurgence - population-level implications of the Israeli campaign

**DOI:** 10.1101/2022.01.18.22269141

**Authors:** Nir Gavish, Rami Yaari, Amit Huppert, Guy Katriel

## Abstract

Israel was one of the first countries to administer mass vaccination. Consequently, it was among the first countries to experience significant breakthrough infections due to the waning of vaccine-induced immunity, which led to a resurgence of the epidemic. To mitigate the outbreak, Israel launched a pioneering booster campaign. Israel’s success in curtailing the Delta resurgence while imposing only mild non-pharmaceutical interventions has been instrumental in influencing the decision of many countries to initiate a booster campaign. By constructing a detailed mathematical model and calibrating it to the Israeli data, we expand our understanding of the impact of the booster campaign from the individual to the population level. We use the calibrated model to explore counterfactual scenarios in which the booster vaccination campaign is altered and to assess the direct and indirect effects in the different scenarios. Our results point to the vast benefits of vaccinating younger age groups that are not at a high risk of developing severe disease but play an important role in transmission. We further show that when the epidemic is exponentially growing the success of the booster campaign is highly sensitive to the timing of its initiation. Hence a rapid response is an important factor in reducing disease burden using booster vaccination.

## 1 Introduction

During June 2021, Israel experienced an exponential rise in COVID-19 cases, with many infections and severe cases reported among vaccinated individuals. At that stage, roughly 80% of the eligible population and two-thirds of the entire population were vaccinated, following a successful campaign using two doses of the BNT162b2 vaccine [1–3]. Initially, it was unclear to what extent resurgence was driven by increased infectiousness of the Delta variant, by heightened immune evasion, or by the waning of vaccine-elicited immunity. A nationwide study in Israel reduced the uncertainty by demonstrating a strong effect of waning immunity in all age groups six months from vaccination [4]. Israel faced a dilemma regarding the administration of booster vaccinations since, at that stage, the BNT162b2 booster vaccination had not yet been approved by the United States food and drug administration (FDA) or any other regulatory agency.

In order to curtail the Delta outbreak, Israel began administering booster vaccinations on July 30, 2021. Initially, booster vaccinations were restricted to ages 60 and older, but eligibility was rapidly extended to other age groups, so that by the end of August 2021, individuals of age 16 and older who were at least five months past their second dose were eligible for booster vaccination. During August, 2.25 million booster vaccinations were administered, and by December 2021, an overall of ∼4 million individuals (∼80% of the eligible population) received the booster.

The example of Israel’s success in curtailing the Delta resurgence without a lockdown and with mild non-pharmaceutical interventions has been instrumental in influencing the decision of many nations to initiate a booster campaign. There is, however, an ongoing debate and uncertainty regarding the required extent of a booster campaign, e.g., which age groups should be boosted, and the importance of a rapid vaccination effort.

In this work, we present an in-depth analysis of the population-level impact of the different elements of the booster campaign on epidemic outcomes. To do so, we develop a transmission model that incorporates the waning of vaccine-induced immunity, and its buildup after boosting. The model accounts for vaccine and booster administration per age group at a daily resolution and is calibrated using real-world data of Israel in the period from July 1^st^ to November 25^th^ 2021. The model is fitted to time series of PCR confirmed cases in 10-year age groups and different vaccination states. The calibrated model is used to study the impact of the booster campaign by quantifying the outcomes of counter-factual scenarios, e.g., the application of alternative boosting campaigns in which boosting is restricted to specific age groups or in which the timing of the booster campaign is modified.

The direct protection afforded to those vaccinated is estimated by conducting clinical trials and observational studies comparing outcomes in vaccinated and unvaccinated individuals [4, 5]. However, the indirect protection provided by reducing transmission at the population level is a key consideration when evaluating vaccination policies. The model is used to disentangle indirect effect by comparing outcomes of the vaccination campaign to expected outcomes in the absence of vaccinations. By modelling the ‘competition’ between the vaccination campaign and the spread of the epidemic, this study thus demonstrates the contribution of mathematical modelling to analyzing data and obtaining a retrospective understanding of the role of the different mechanisms in generating the observations. Such an understanding is crucial for making future projections and planning interventions.

## 2 Methods

In order to capture the transmission dynamics of the Israeli fourth wave, we developed a discrete-time age-of-infection age-stratified transmission model, which includes vaccination and booster administration. We use nationwide high-resolution data from Israel to calibrate and validate the model. A complete account of the data, model, and methods used is provided in the supplementary information. Here we provide a brief description of the main details.

### Data

The data was extracted from the Israel Ministry of Health’s database. The information per case consists of age, place of residency, dates of PCR tests, vaccination dates (first, second, and third doses), COVID-19 related hospitalization (mild or severe disease), and mortality. Severe disease was defined according to the National Institutes of Health COVID-19 treatment guidelines [6].

### Waning rates

We rely on the estimates of the waning rate of vaccine protection from infection in [4, 7] to estimate the loss in protection as a function of time since vaccination. The waning rate of vaccine protection from severe outcomes is assumed to be equal to the waning rate of vaccine protection from infection. This assumption is validated using the relative estimates in [4]. See SM for additional details.

In the absence of data supporting quantitative estimates, we assume that the waning rate of booster protection from infection and severe outcomes is half the waning rate of second dose vaccine protection from infection and severe outcomes.

### Transmission model and parameter inference

We developed a discrete-time age-of-infection age-stratified transmission model. The infection process is modeled using a social contact matrix which determines the average number of daily interactions, e.g., the number of contacts a 53-year-old has with individuals in the 30-39 age group. The social contact matrix is composed of a linear combination of contact matrices customized to Israel in key social settings [8]: household, work, community, and school. The relative weight of the school matrix is set according to school vacations, whereas the relative weights of the other three matrices are determined using Google’s COVID-19 community mobility data [9] (see SM for details). Additional parameters are inferred by fitting the model to data collected during the period from July 1^st^ to November 25^th^ 2021, in which the Delta surge took place in Israel. Specifically, we consider the time series of confirmed cases stratified by 10-year age groups and vaccination status. These amount to an overall 27 time series at daily resolution. We used these time series to estimate four parameters using maximum likelihood: the transmission parameter, a parameter corresponding to the relative susceptibility of children, and two parameters related to the age-dependent detection rate of individuals.

We observe that the model agrees with the time series of severe cases stratified, as the detection data, to 27 time series at daily resolution (see SM).

## 3 Results

### Capturing the dynamics of the Delta outbreak

The development of a model calibrated by real-world data of Israel during the Delta surge is a key outcome of this work, as it provides a basis for the study. Figure 1 demonstrates that the model captures the dynamics well, both before and during the period of the booster vaccination campaign. It relies on only four fitted parameters, see methods and SM for details. More so, all estimated parameter values are within reasonable epidemiological values, a fact which further confirms the appropriateness of the modeling scheme. We note that, during September, the epidemiological data shows some from variations and irregularities primarily due to a sequence of Jewish holidays, which affect the level of the testing effort as well as patterns of transmission.

**Figure 1:**
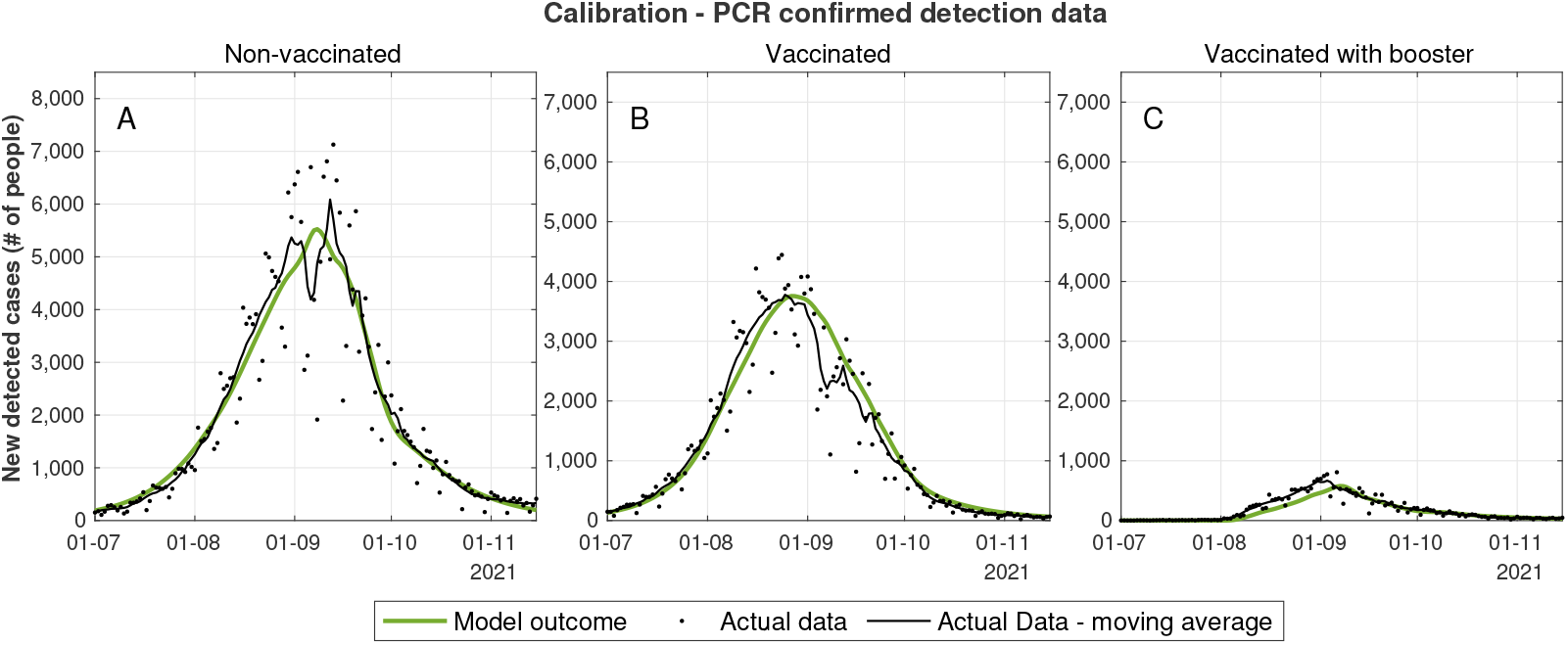
Calibrated model outcomes vs actual data. Daily number of new detected cases who where not vaccinated (A), who received two vaccine doses (B) and those who received three vaccine doses (C). The data series presented in these graphs are aggregated from 27 data series which were used for model calibration. Graphs corresponding to each of the data series are presented in the SM.

Following calibration of the model, we first use it to explore the scenario in which *no booster vaccinations* are administered, and no additional non-pharmaceutical interventions are implemented. In the absence of boosters, the model projects the continued rise of a significant outbreak reaching a peak of ∼ 37,300 cases detected daily and ∼ 770 new severe cases per day with a significant portion of individuals who had received two doses of vaccine, see red dashed curves in Figure 2. These numbers of severe cases are far beyond the threshold at which medical care in Israel can be provided without being severely compromised. Indeed, Israel reached a peak of roughly 190 new severe cases per day in previous surges. Such patient loads in hospitals had been estimated to have resulted in as much as 25% excess in-hospital mortality in Israel [10]. In fact, during previous surges, severe non-pharmaceutical interventions were applied at much lower rates of severe cases. For example, Israel entered a lockdown on September 18th, 2020 with ∼80 new severe cases per day and to an additional lockdown on December 27th, 2020 with ∼100 new severe cases per day.

**Figure 2:**
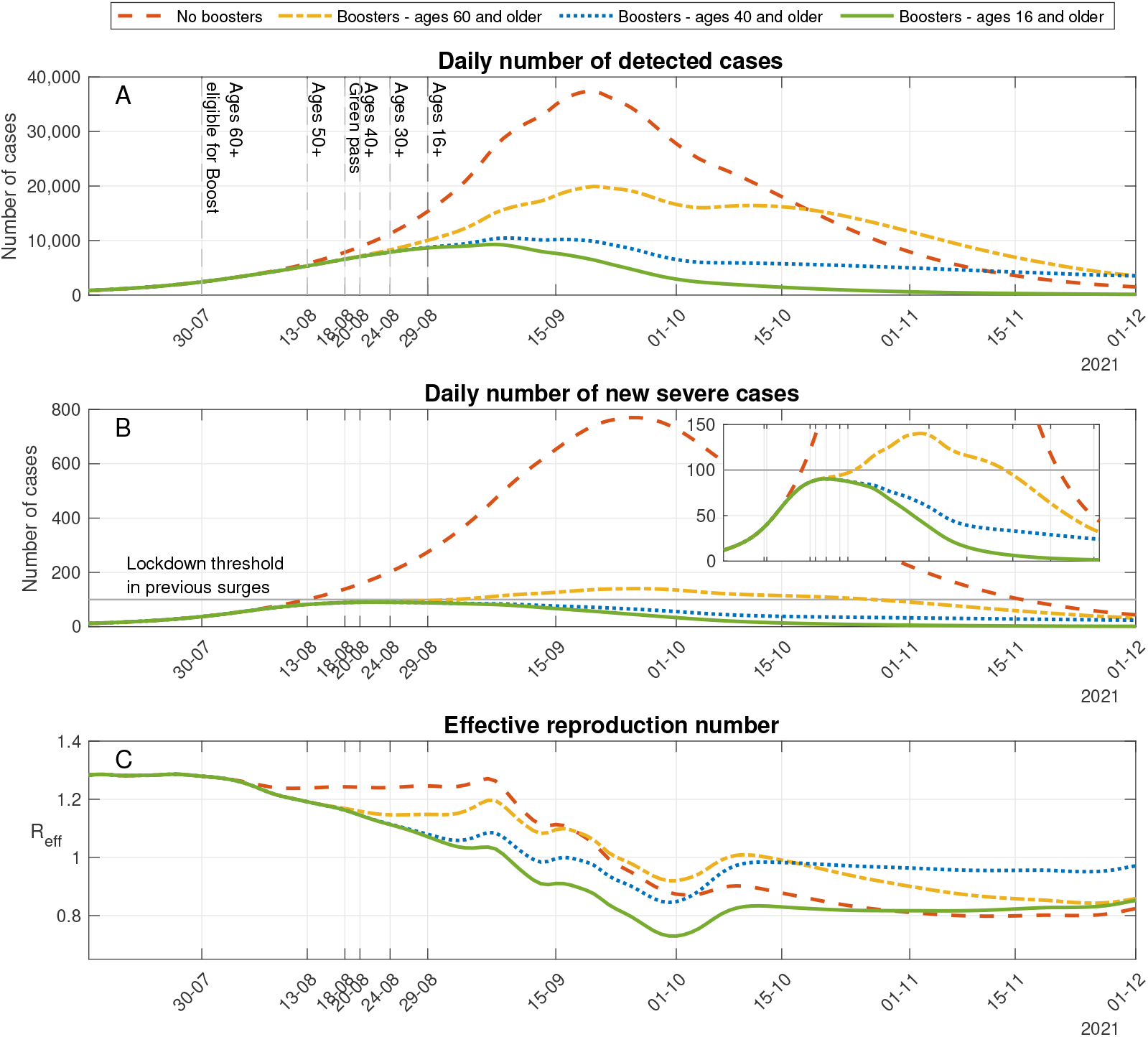
Effect of waning and boosting. Model projections for epidemic spread in a scenario in which no boosters are administrated (**Dashed curve**), boosters are administrated only for those of age 60 and older (**Dash-dotted curve**), boosters are administrated only for those of age 40 and older (**Dotted curve**) and a scenario which accounts for the actual numbers of boosters that were administered until the end of November 2021 (**Solid curve**). A: Daily number of detected cases. B: Daily number of new severe cases. Inset graph presents the same data as in B, but with a zoom-in on lower values of the y-axis. C: Effective reproduction number.

The assessment of the situation during July indicated the urgent need of applying non-pharmaceutical interventions or other restrictions to prevent catastrophic outcomes. In parallel, intensive research efforts led to the understanding that the waning of vaccine efficacy in Israel is a dominant factor in the resurgence of the epidemic spread [4]. Additionally, using the same modeling framework presented in this work, quantitative assessment of the population-level effects of administrating a booster vaccine to the vaccinated population, particularly those vaccinated at the initial stages of the Israeli vaccination campaign, showed significant benefits in most scenarios tested, with a sharp reduction in the number of cases and the burden of hospitalizations. Following these studies, and the recommendation of the pandemic advisory committe, the Israeli ministry of health decided to launch a booster vaccination campaign.

### Curtailing the outbreak using boosters

On July 12, 2021 Israeli health authorities recommended the administration of a third dose, to high-risk populations. The official booster campaign was initiated on July 30^st^ with a recommendation to vaccinate those of age 60 and older who had received a second Pfizer vaccine dose at least five month before. The age of eligibility was gradually expanded during August: Ages 50-59, August 13; Ages 40-49, August 20; Ages 30-39 on August 24, and ages 16 and older on August 29. During the campaign, ∼42% of the population, 4 million individuals, were given a booster vaccination (see [5] for details). To assess the population-level effect of the actual booster campaign in Israel, we rely on the model calibrated with data from the Delta surge in Israel (see Figure 1) and compare the model outcomes to model projections in a ‘hypothetical’ scenario in which no booster vaccinations are administered.

Comparison of the red dashed line (No boosters) with the solid green line (Booster for ages 16+) in Figure 2 demonstrates the dramatic impact of the booster campaign in curtailing the outbreak. In particular, we find that the overall number of cases in the simulation that accounts for booster uptakes is 74% (CI; 73.9%-74.3%) smaller than the number of cases projected during the resurgent wave in the absence of boosters or other interventions. Similarly, we estimate that booster vaccination reduced mortality by 88.6% (CI; 88.5%-89%).

### Assessing different boosting policies

Israel chose to administer the booster to the general population rather than concentrate on the elderly (ages 60 and older) and other high-risk groups. There is an ongoing debate as to whether such a policy is best suited when the primary goal is to reduce the load on the health care system. The dispute concerns the magnitude of reduction in severe disease cases due to decrease in transmission rates in low-risk populations. To estimate the impact of boosting the low-risk groups, we consider counter-factual scenarios in which the booster vaccination campaign is modified so that smaller segments of the population are boosted. Specifically, we examine cases in which booster administration is restricted to ages 60 and older or to ages 40 and older. To do so, we repeat the simulation presented in Figure 2 while removing all boosters given to individuals under the age of 60 or 40, respectively, and leaving the rates of vaccination in the eligible groups unchanged.

In the case booster vaccines are given only to age groups 60 and older, we observe that the outbreak is considerably reduced compared to the case in which no boosters are given (see dash-dotted yellow curves in Figure 2). Roughly a third of these averted cases are of ages below 60, demonstrating strong indirect effects of the booster vaccination (see below for a quantitative assessment of the indirect effect of the booster). The outbreak, however, is not reduced to the same extent as in the case boosters are provided to all eligible ages (green solid line). Indeed, we observed that, under the conditions considered, restricting booster eligibility to ages 60 and older leads to an increase of 303%, 232% and 209% in the number of confirmed, severe cases and deaths respectively, relative to the number of cases when all eligible age groups are provided a booster dose (see Table 1).

**Table 1:** Change in epidemiological outcomes in various booster policies relative to the outcomes of the actual booster campaign.

| Booster Policy | Change in cases | Change in severe cases | Change in mortality |
| --- | --- | --- | --- |
| No Boost | 385% (384-390) | 763% (757-783) | 880% (871-905) |
| Ages 60 and above | 303% (302-306) | 232% (231-236) | 209% (207-214) |
| Ages 40 and above | 184% (184-186) | 146% (145-149) | 144% (143-148) |
| Early schedule | 56.9% (56.7-57.4) | 53% (52.8-54) | 52% (51.5-53.3) |
| Late schedule | 162% (162-164) | 178% (177-182) | 184% (182-188) |

As expected, when boosters are restricted to age groups 40 and older, the outcomes are closer to those attained when boosters are given to ages 16 and older. Nevertheless, there is a significant increase of 184%, 146% and 144% in the number of confirmed, severe cases and deaths respectively (see Table 1 and dotted blue curves in Figure 2). An important difference between the outcomes is that in the case boosters are given to ages 40 and older, the epidemic slowly decays until an effective reproduction number of roughly 0.9 is reached (see Figure 2C). This slow decay leads to a projected average of 3,600 confirmed cases and 30 new severe cases per day in late November. In comparison, when boosters are given to ages 16 and older the decay of the epidemic is faster with an effective reproduction number of ∼0.825, leading to 350-400 confirmed cases and 3-4 new severe cases per day in late November (see Figure 2C). Consequently, when the booster campaign curtails the epidemic spread, the risk of a resurgence due to a new variant, seasonality, national holidays, or future waning increases when booster eligibility is restricted to a smaller population. The reason is that fewer people gain protection through vaccination or infection.

### Timing of the boosting campaign

The Israeli Ministry of Health debated the need for a booster campaign during the entire month of July. In early July, it was unclear to what extent the resurgence was driven by increased infectiousness of the Delta variant, by heightened immune evasion, or by the waning of vaccine-elicited immunity. Accordingly, the potential effectiveness of administering boosters in curtailing the outbreak was also unclear. The picture was partially clarified in mid-July, as it became evident that waning vaccine immunity plays a crucial role in driving the outbreak. Israeli authorities decided to initiate an extensive booster campaign within two weeks of this observation. It is therefore interesting to evaluate potential outcomes in case the booster campaign had been initiated at an earlier date, as well as the consequences had the decision to initiate the campaign had been somewhat delayed. We address these question by running the model simulation with the calibrated parameters, changing only the starting date of the vaccination campaign.

Figure 3 shows that delaying or advancing the initiation of the vaccination campaign by two weeks leads to major differences in the outcomes of the booster campaign. As expected, delaying the booster vaccination campaign by two weeks results in a larger outbreak (see for comparison the differences between the solid blue line and the dashed yellow curve in Figure 3). In this case, we find that the peak number of severe cases and mortality are comparable to those attained when boosters are restricted to ages 60 and above. In the reverse case, in which the booster vaccination campaign is advanced by two weeks, the epidemic spread is curtailed at an early stage. The early epidemic decay leads to a reduction of 57%, 53% and 52% in the number of confirmed, severe cases and deaths, respectively, relative to the numbers attained with the actual booster schedule, see the differences between the solid blue line and the dash dotted red curve in Figure 3 and Table 1. In this case, however, the epidemic decay is very slow. Eventually, by mid-October it stagnates with an effective reproduction number slightly below one. Accordingly, the projected number of cases in late November is 1,300 confirmed cases and 10 new severe cases daily. In comparison, the model projections corresponding to the actual booster campaign give rise to roughly 3 times less confirmed and severe daily cases in the same period. Consequently, we observe that the early administration of booster vaccines efficiently curtails the epidemic wave in the short term. However, it increases the risk of resurgence in the longer run due to the fact that fewer recovered individuals are accumulated.

**Figure 3:**
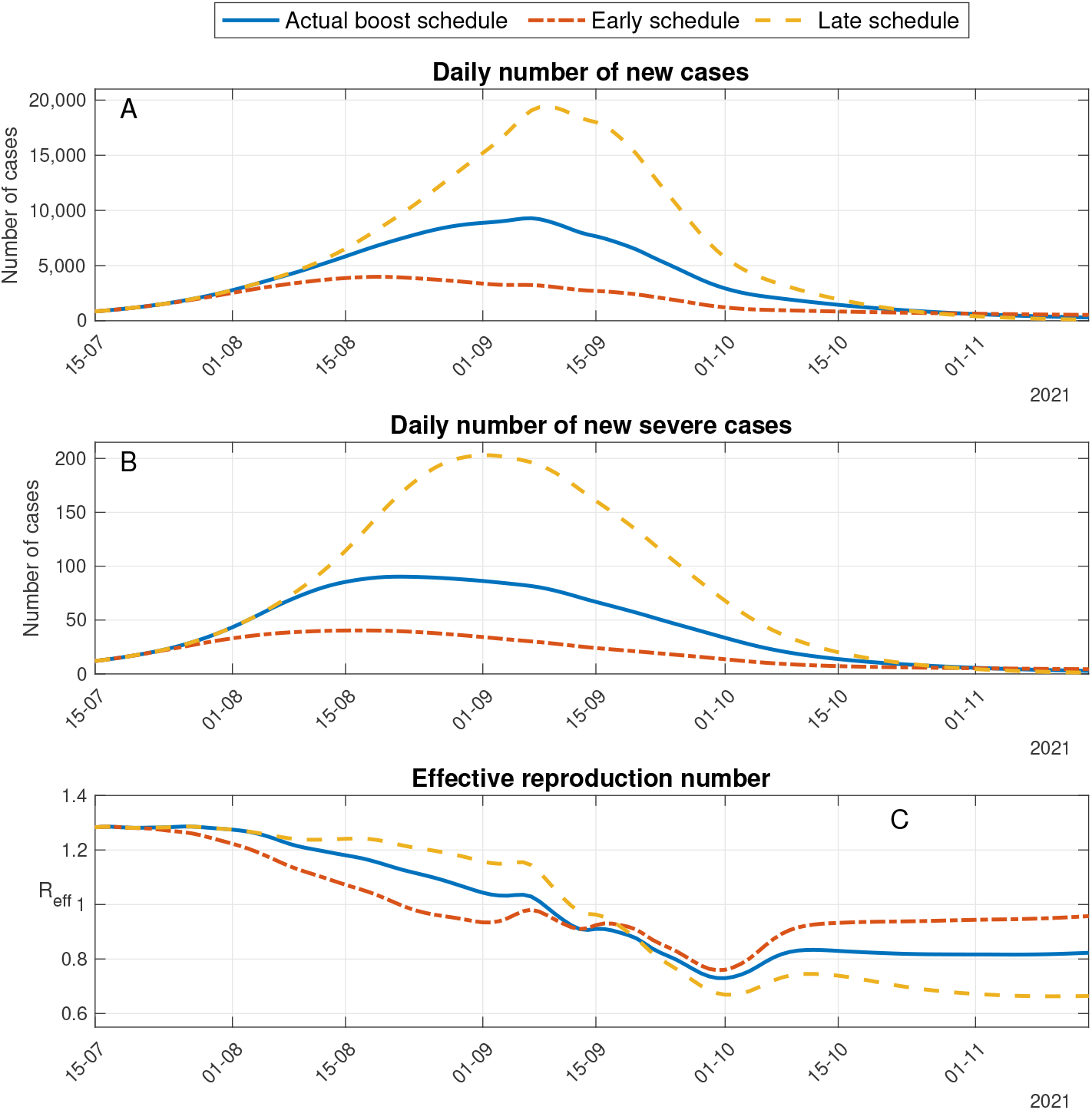
Timing of boost start date. Model projections for epidemic spread in a scenario in which the booster campaign had been delayed (**Dashed curve**) or advanced by two weeks (**Dash-dotted curve**), compared to a scenario which accounts for the actual timing of the booster campaign (**Solid curve**). A: Daily number of detected people. B: Daily number of new severe cases. C: Effective reproduction number.

### Quantification of direct and indirect effects

Indirect benefits are a crucial consideration when evaluating SARS-CoV-2 vaccination policies [11], but disentangling indirect effects from numerous types of epidemic field observations is challenging [12]. Here, we follow [13] and use simulations to explicitly quantify the indirect protective effect of the booster vaccination campaign, due to reduction of transmission. To do so, we use the model to compute epidemic outcomes in a scenario in which booster vaccination is regarded to be non-protective. In such a case, the direct effect of booster vaccination is measured as the number of infections among those who received the booster, since these infections would not have occurred if the booster vaccine were fully protective. The indirect effect is computed as the difference between the total excess infections that would have occurred if boosters were not administrated and the number of infections prevented by the direct protective effect [13]. Our computation shows that among the cases reduced due to the booster campaign, ∼55% were reduced due to direct protection, while the rest are due to indirect protection. We further find that ∼82% of the new daily severe cases are reduced due to direct protection, while the rest are due to indirect protection. We note that the above computation does not take into account the occurrence of breakthrough infections among those who received a booster. The very low rate of such breakthrough infections in the relevant period justifies this approximation. Accounting for breakthrough infections in the computation would give slightly more weight to indirect protection.

The above results quantify the significant impact of booster vaccination in reducing transmission and providing indirect protection to those susceptible to infection. In particular, these results points to the vast benefits of vaccinating younger age groups that are not at a high risk of developing severe disease but do play an important role in transmission.

## 4 Discussion

Israel was one of the first countries to administer mass vaccination and execute it with very high rollout rates. Due to the early vaccination, Israel was also among the first countries where the waning of vaccine protection occurred. Identification of the nature of vaccine waning [4] led Israeli authorities to pioneer a booster campaign that curtailed the Delta resurgence. Previous studies have quantified the effects of boosting on an individual level [5]. Here, we expand our understanding to the population level by constructing a mathematical model that accurately describes the Delta surge dynamics in Israel. The modeling framework allows us to compare alternative boosting policies to ascertain the trade-offs of vaccinating different age groups and the importance of an early and extensive response. In addition, the mathematical model allows one to explore the relative role of direct and indirect protection on different outcomes of the disease.

Our results indicate that, if a booster campaign had not been undertaken, Israel would have needed to apply extensive non-pharmaceutical interventions to prevent a destructive epidemic wave. To quantify the potential severity of the Delta outbreak, we used a counterfactual assumption that boosters had not been provided. The model estimated that without boosting or non-pharmaceutical interventions, the potential of the Delta wave was an increase of ∼880% in mortality, of ∼763% in severe disease, and a ∼385% increase in detected infections. Overall, providing a booster vaccination to roughly 40% of the population led to a ∼70% decrease in the number of cases. The gap between these numbers demonstrates the significant indirect protection of the booster vaccination.

Quantifying the extent of direct and indirect protection is crucial in building strategies to cope with the pandemic or end it [11]. The direct protection of the booster has been estimated in both clinical trials and observational studies [5, 14, 15]. Indirect protection was estimated in a household study [16], and found that the odds ratio of a secondary case in a vaccinated individual was 0.54 of an unvaccinated person. However, the full extent of the indirect protection by reducing transmission can not be fully estimated in the field. Mechanistic modelling can help bridge this gap. Indeed, we demonstrate the use of the model to explicitly quantify the indirect protective effect of the booster vaccination campaign by studying appropriate counterfactual scenarios.

In addition, we assess the expected impact of more restricted booster vaccination campaigns and compare alternative policies. Restriction of booster availability to age groups 40+ or 60+ would have significantly reduced infection, severe disease, and deaths (see Table 1). On the other hand, it is far less effective in all the outcomes than boosting more segments of the population. We further show that when the epidemic is exponentially growing, the success of the booster campaign is sensitive to the timing of the initiation. For instance, advancing the campaign two weeks earlier would have led to an overall decrease in the number of cases by roughly a factor of three compared to the actual booster campaign. This sensitivity to the timing manifests the competition between the epidemic spread and the booster rollout rate and the fact that booster vaccinations provide excellent protection to an individual within a short period from receiving the vaccination.

We have used the data and parameter values representing the population’s epidemiology and behavior during the Delta resurgence in Israel to calibrate the model. The focus of this case study raises a natural question regarding the extent to which our conclusions can be generalized. The transmission dynamics are sensitive to the social contact patterns. The social contact matrix used in the model is composed of a time-varying linear combination of contact matrices in key social settings [8] which is determined using Google’s COVID-19 community mobility data. We found that the number of social contacts of those above the age of 60 during the Delta outbreak in Israel was relatively high compared to previous waves and the number of social contacts described by the commonly used POLYMOD contact matrix in this age group. Indeed, the contribution of age group 60 and older to the basic reproduction number in the time-varying social contact matrix used in the simulation varies about 10-11%, while the contribution of age group 60 and older to the basic reproduction number in the POLYMOD contact matrix adapted to Israel [17] is just 2%. We also note that the impact of extending eligibility for the booster vaccine to younger age groups also crucially depends on the vaccine uptake rates, which is lower in younger age groups (see Figure 6 in the SM). The above considerations warrant care in adapting the results to other countries.

This work aims at understanding the population-level effect of booster vaccine protection at the shortterm. Accordingly, the model neglects factors that are likely to impact epidemic outcomes in the coming years. In particular, we did not account for demographic turnover (births, deaths, and aging), waning of convalescent immunity, or seasonality in the transmission rate. To conclude, this study highlights the importance of using boosters to curtail outbreaks resulting from waning immunity. The modeling demonstrates that both a rapid response, and one that includes the sub-populations which plays a significant role in transmission, even if they are at low risk of severe disease, are significant for reducing the disease burden. As the world faces the danger of new variants of concern, such as the Omicron threat, it is crucial to improve our ability to minimize the toll of the disease.

## Data Availability

All data and source codes used in this study are avaliable online at https://github.com/NGavish/boosterVaccinationModel.git

## Ethics statement

The study was approved by the Institutional Review Board of the Sheba Medical Center. Helsinki approval number: SMC-8228-21.

## Supplementary Materials - Materials and Methods

### A Data sources

The incidence data sets include all PCR confirmed (detected) and severe Covid-19 cases in Israel between July 1^st^ to November 25^th^ 2021. The data sets are stratified by age-group (9 age-groups: 0-9,10-19,20-29,30-39,40-49,50-59,60-69,70-79,80+) and by vaccination status: unvaccinated, vaccinated with two doses and booster-vaccinated.

The data files are available at github.com/NGavish/boosterVaccinationModel.git.

Note: Data cells with non-zero values smaller than 5 were randomized to comply with the Israeli Ministry’s of Health regulations.

### B Transmission model

We develop a discrete-time age-of-infection model (also known as renewal equation-type model). Accounting for age-of-infection enables us to take into account the appropriate generation-time distribution. Additionally, it enables us to precisely incorporate the processes of primary and booster vaccination based on the detailed data at daily resolution of the vaccination schedule, and the increase of susceptibility of those vaccinated with time since vaccination, as estimated in previous studies from Israel.

The source code of the transmission model is available at github.com/NGavish/boosterVaccinationModel.git. In what follows, we describe the main components and computations of the model.

#### Dynamic variables

The population is divided into *n* = 9 age groups. The discrete variable *t* refers to time in days, where *t* = 0 is the first day on which vaccinations were administered in Israel (Dec. 20,2020). We distinguish between non-vaccinated individuals (nv), vaccinated (v) and those vaccinated with three doses (b), and compute the evolution of the following variables:

- *S*(*t, j*) is the overall number of susceptible, non-vaccinated individuals of age group *j* on day *t*.
- *V* (*t, j, s*) is the overall number of individuals of age group *j* who had been vaccinated *s* days before day *t*, and have not been infected up to day *t*.
- *B*(*t, j, s*) is the overall number of individuals of age group *j*, who have received a booster dose *s* days before day *t*, and have not been infected up to day *t*.
- *i*_*nv*_(*t, j*) is the number of newly infected individuals of age group *j* on day *t* who have never been vaccinated.
- *i*_*v*_(*t, j, s*) is the number of newly infected individuals of age group *j* on day *t* who were vaccinated *s* days before day *t*.
- *i*_*b*_(*t, j, s*) is the number of newly infected individuals of age group *j* who received a booster vaccination *s* days before day *t*.

#### Model equations

The daily incidence of infections of unvaccinated individuals in age group *j* at day *t* is given by

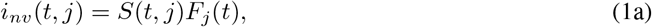

where *F*_*j*_(*t*) is the force of infection or the daily probability of an unvaccinated individual from age group *j* becoming infected. The force of infection is given by

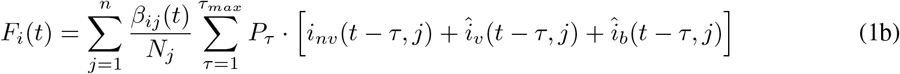

where 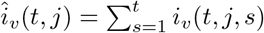 and 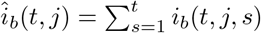 are the total number of individuals of age group *j* who have received second shot or booster vaccination, respectively, and become infected on day *t*. Additionally, *P*_*τ*_ (1 *≤ τ ≤ τ*_*max*_, here *τ*_*max*_ = 14 days) is the generation time probability density function, *β*(*t*) is the time-varying transmission matrix whose *ij*^*th*^ element denotes the transmission rate between an individual from age-group *j* and individuals of age-group *i*; See below for details on the estimation of the generation-time distribution and the construction of the transmission matrix.

Similarly, the daily incidence of infections of vaccinated individuals in age group *j* at day *t* is given by and

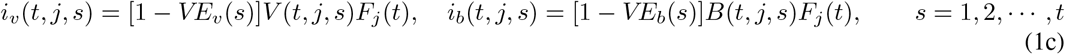

where *VE*_*v*_(*s*) and *VE*_*b*_(*s*) is the profile of vaccine protection against infection of individuals *s* days after first dose vaccination or booster vaccination, respectively.

The simulation progresses day by day, *t* = *t*_0_, *t*_0_ + 1, *· · ·*, where *t*_0_ is the start time of the simulation (*t*_0_ = 194 in this work, corresponding to the time from the first vaccination in Israel to July 1,2021, the start of our simulation). For each day *t* and group *j*, it performs the following steps:

1. Compute the daily incidence of infections, using (1).
2. Update the number of susceptible individuals, taking into account infections and vaccinations with the first dose.

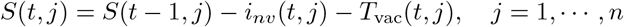

where *T*_vac_(*t, j*) is the number of individuals of age group *j* who are vaccinated on day *t*.
3. Update number of vaccinated, taking into account infections, first dose vaccinations and booster vaccination

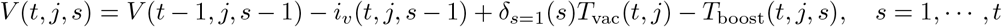

where *T*_boost_(*t, j, s*) is the number of individuals of age group *j* who were vaccinated *s* days before day *t* and received a booster vaccination which became effective on day *t*, and *δ*_*s*=1_(*s*) = 1 when *s* = 1, and zero otherwise.
4. Update number individuals who have received the booster, taking into account infections, and new booster vaccinations

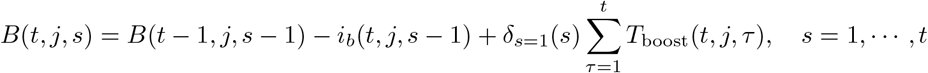

#### Model parameters

Table 2 summarizes the list of model parameters and the basis for setting their values. In addition, the model inputs includes the transmission matrix and vaccination schedules. In what follows, we provide additional details on the parameters and inputs.

**Table 2:**
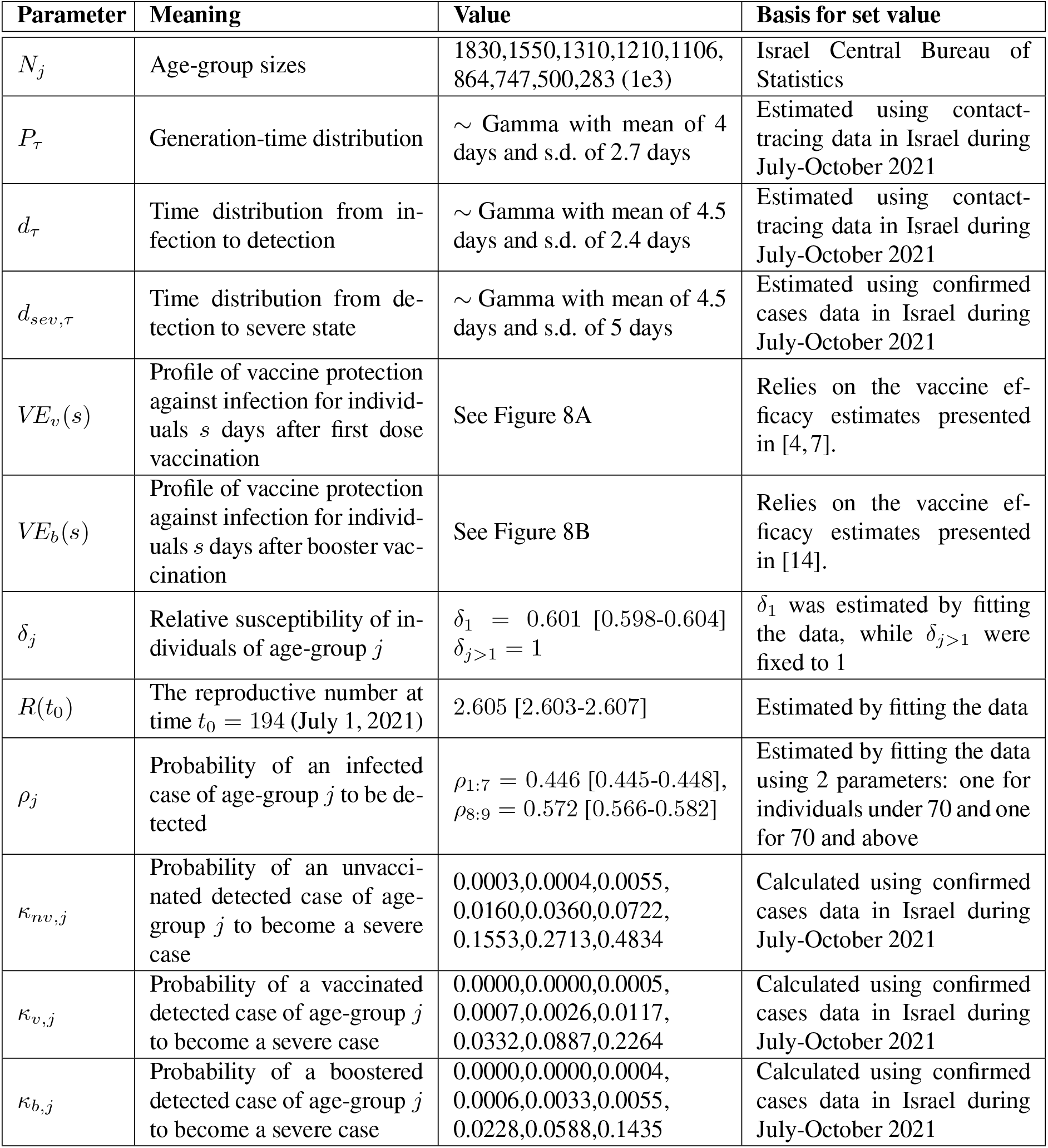
Model parameters.

##### Generation time distribution

The generation time distribution *P*_*τ*_ was set to a discretized version of a gamma distribution with a mean of 4 days and a standard deviation of 2.7 days, based on intervals between infections obtained from over 5000 triplets of known infector-infectee/infector-infectee in the data set of confirmed cases in Israel during July-October 2021.

##### Transmission matrix

The transmission matrix *β* is given by

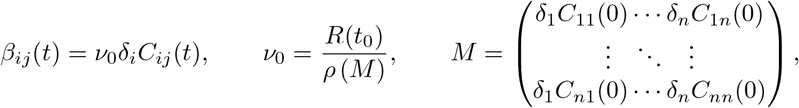

where *C* is an age-group contact matrix, *δ* is a vector containing the relative susceptibilities of each age-group (set to 1 except for the young children age-group 0-9 whose relative susceptibility was obtained by fitting the data), *R*(*t*_0_) is the basic reproductive number at time *t* = *t*_0_ (that is the reproductive number in the absence of immunity induced by vaccinations and previous infections), and *ρ*(*M*) is the spectral radius of a matrix *M*.

We modeled the contact matrix *C* as follows:

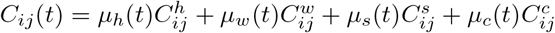

where *C*^*h*^, *C*^*w*^, *C*^*s*^, *C*^*c*^ are household/work/school/community contact matrices, respectively, derived for Israel using census and survey data on key socio-demographic features [8] (see Figure 4). The coefficients *μ*_*h*_(*t*), *μ*_*w*_(*t*), *μ*_*s*_(*t*) and *μ*_*c*_(*t*) measure the contribution of contacts within the different settings to the overall contacts at time *t*. The coefficients for households, workplaces and community contacts are obtained from Google’s COVID-19 community mobility report for Israel during the modeled period [9]. The school coefficients were determined according to the assessed level of operation of formal and informal educational institutions during the modeled period. In particular, this period included both summer vacation during July-August and Jewish holidays during September (see Figure 5).

**Figure 4:**
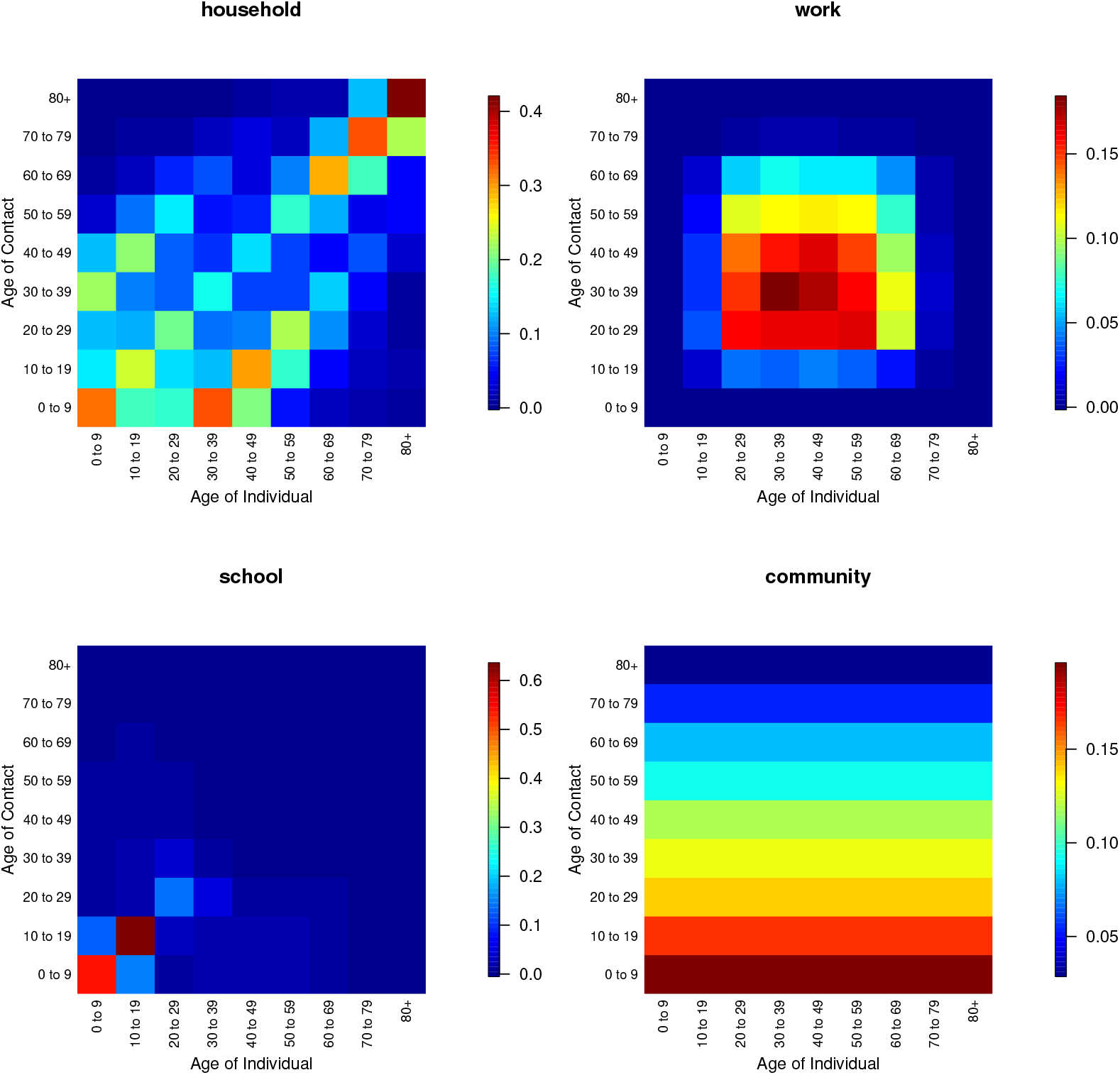
Contact matrices used in the fitting procedure (taken from [8]).

**Figure 5:**
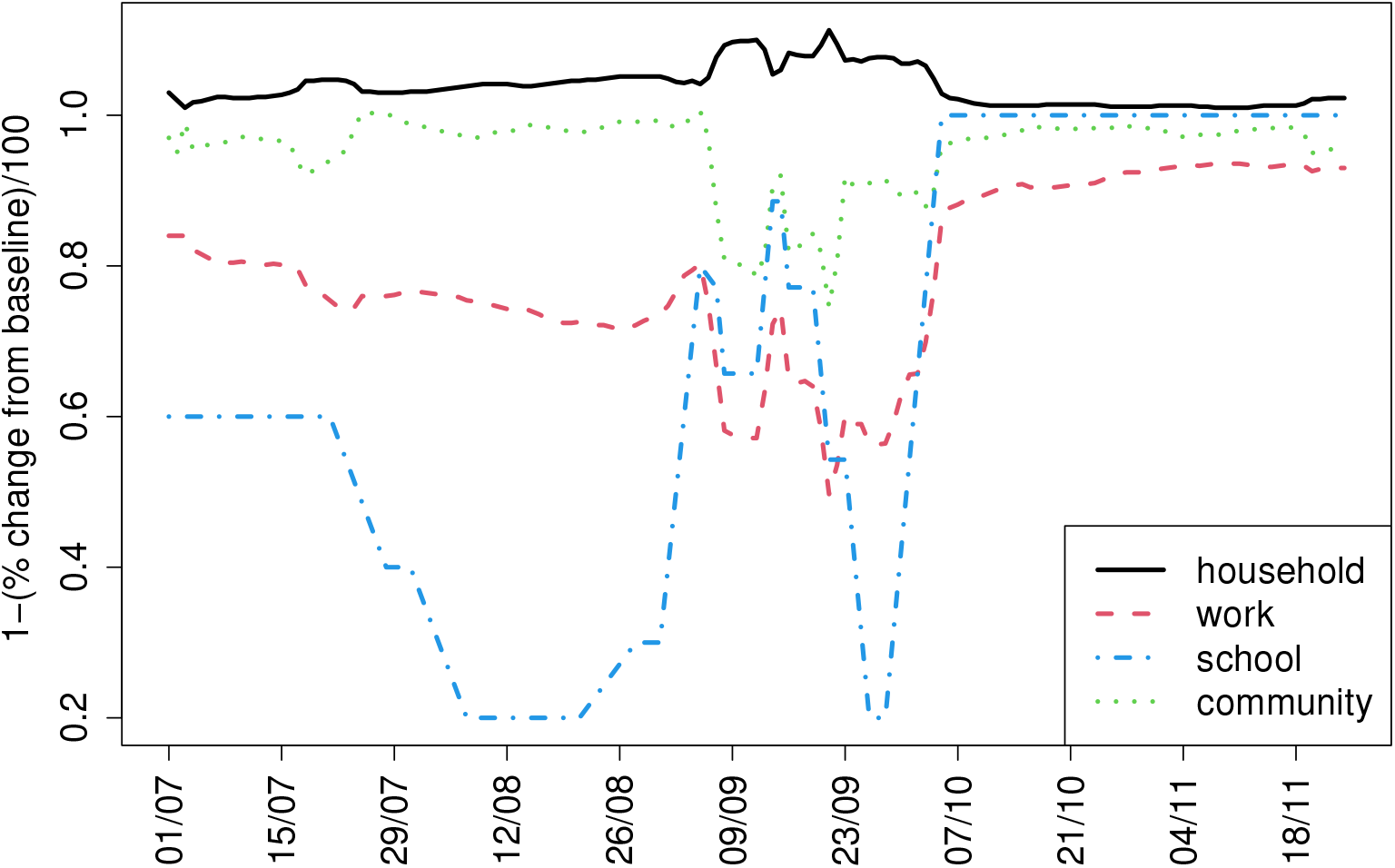
Daily coefficients for the contact matrices used in the fitting procedure. The *household* coefficient is based on Google Mobility Report *residential percent change from baseline*. The *work* coefficient is based on Google Mobility Report’s *workplaces percent change from baseline*. The *community* coefficient is based on Google Mobility Report’s *retail and recreation percent change from baseline*. The *school* coefficient was set according to the assessed level of school openings at each period. All curves were smoothed using a 7-day moving-average.

##### Vaccination and booster schedules

The vaccination schedule *T*_vac_(*t, j*) includes age-stratified first dose vaccine uptakes at daily resolution, and is acquired from the Israel Ministry of Health’s database. Similarly, we acquire from this database the booster schedule *T*_boost_(*t, j*) which describes the number of individuals of age group *j* who received a booster vaccination on day *t*. Figure 6 presents the booster uptake in Israel. The model, however, requires an extend schedule *T*_boost_(*t, j, s*) which also contains the number of days *s* from the day of receiving the first vaccine shot to the day of receiving the booster. However, the organization of Israel Ministry of Health database makes it cumbersome to extract the distribution of days *s* since first dose vaccination for those who received a booster vaccination on day *t*. We have therefore assumed that those who first who received a first dose of vaccine are also the first to receive a booster vaccine.

Both vaccination schedules are processed to account for vaccination of recovered individuals who were not detected. Particularly, we estimate the number of recovered undetected cases per age groups using the detection rate *ρ*_*j*_, and subtract them from the vaccination numbers in the vaccination schedule.

**Figure 6:**
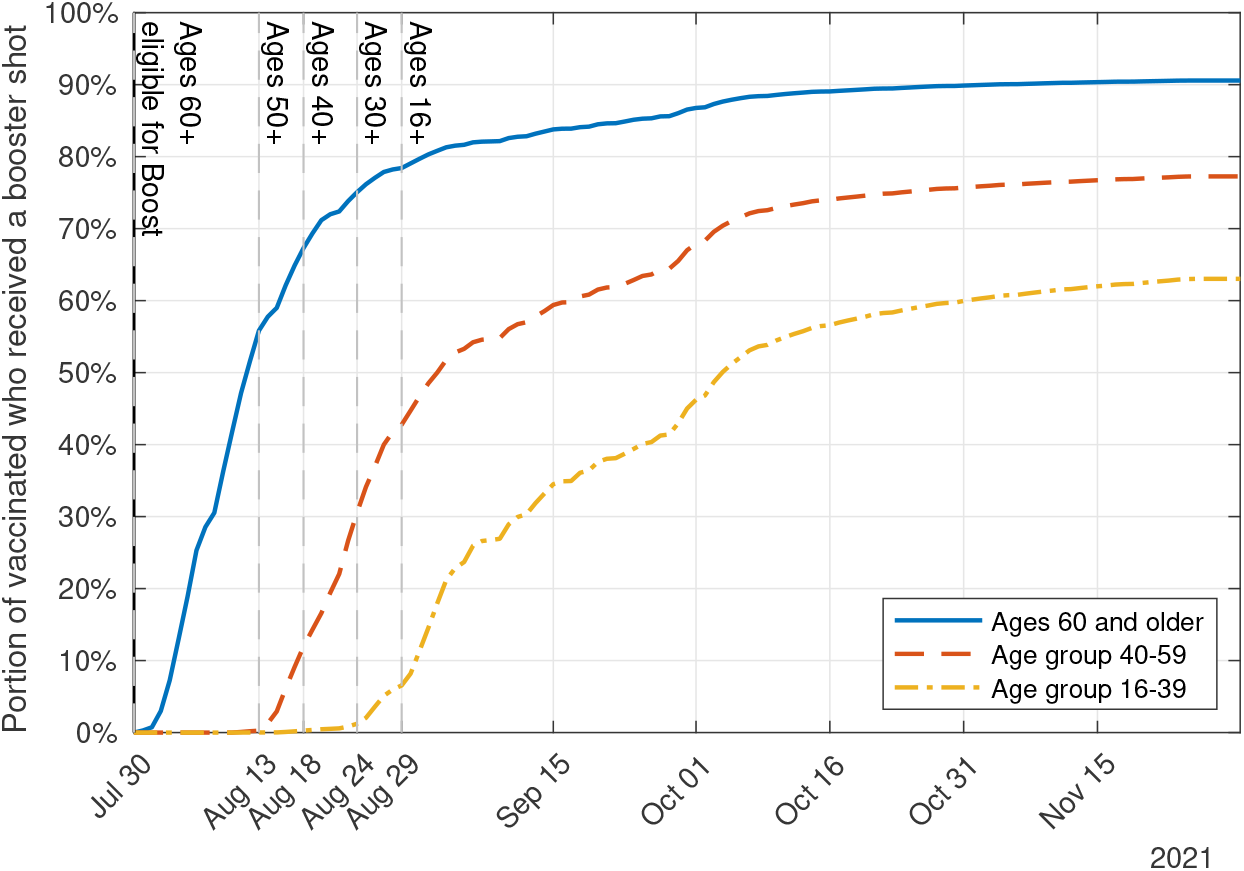
Booster uptake in Israel. The cumulative portion of the vaccinated population in each age group who received a booster shot.

We have verified that the model outcomes are insensitive to the assumptions above.

##### Vaccine efficacy profiles

We rely on vaccine efficacy estimates acquired in a large integrated health system in the USA [7], together with recent estimates acquired in a nationwide study from Israel [18] which extends [4] to account for data collected in July-September 2021 (see Figure 7A). We assume the same vaccine efficacy waning profile for all ages, in accordance with studies showing that the efficacy of mRNA vaccines in reducing infections is nearly independent of age [1, 4, 7, 19]. In lack of data on vaccine efficacy in the long term, we further assume that, in accordance with the trend line fitted in Figure 7A., vaccine protection against infection decays to zero roughly 300 days after the vaccination. We have verified that the model outcomes are insensitive to this assumption.

**Figure 7:**
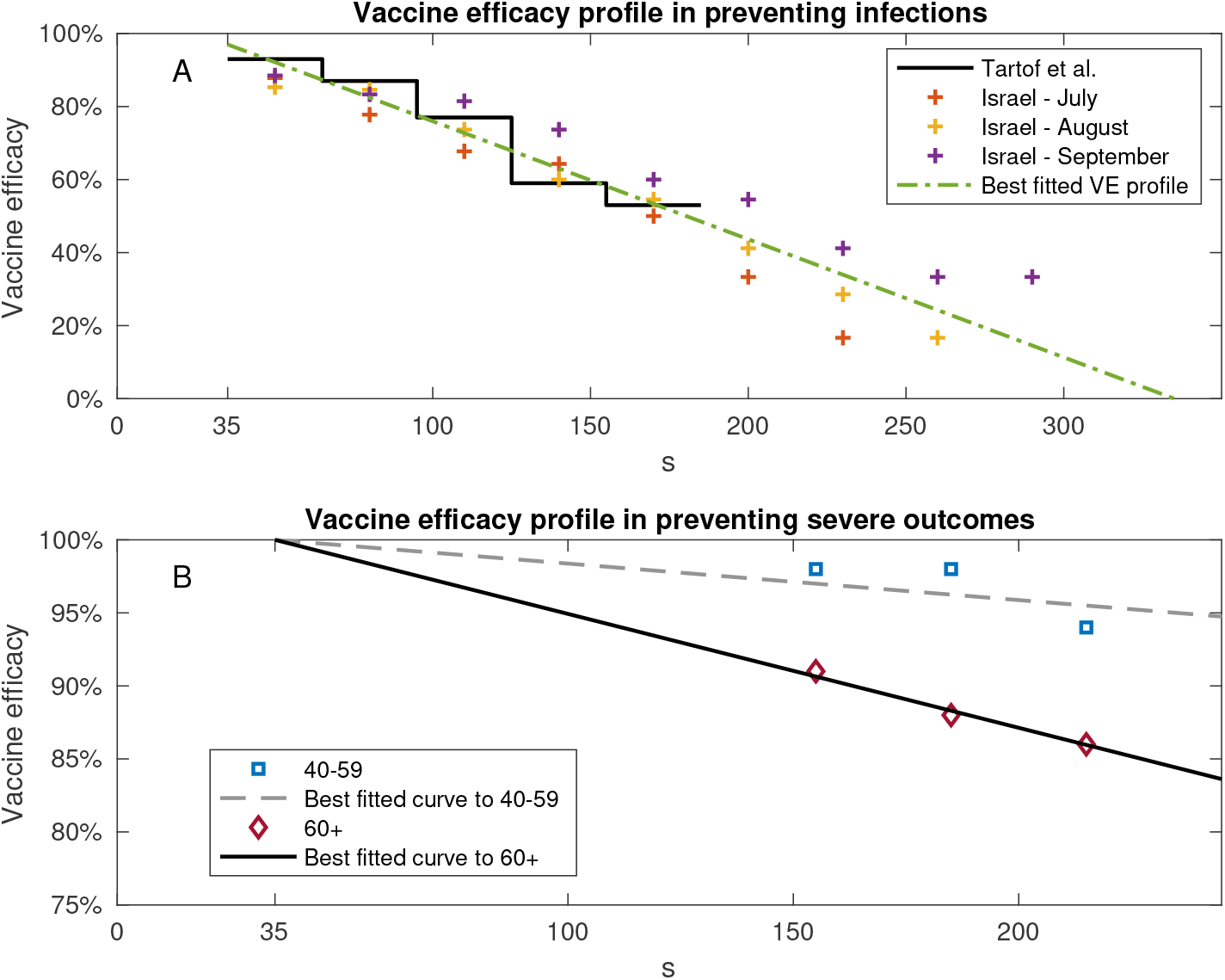
Estimates of vaccine waning profile. A: Estimates of vaccine efficacy profile in preventing infections [7] (Black solid curve), Extension of [4] to account for data collected in July-September 2021 [18] (Cross markers). Superimposed is the best fitted vaccine efficacy profile. B: Estimates of vaccine efficacy against severe outcomes as a function of days after vaccination [4] for ages 40-59 (square blue markers) and ages 60 and older (diamond red markers). Superimposed are the best fitted vaccine efficacy profiles per age groups under the assumption that vaccine efficacy in preventing severe disease wanes at the same rate as vaccine efficacy in preventing infections. In all cases, estimates start from time of full vaccination at day *s* = 35 after receiving first shot.

The short-term booster protection estimated in a nationwide study from Israel [14] is used to extract the efficacy profile *V E*_*b*_(*s*) of individuals *s* days after receiving the booster for *s ≤* 35. In the absence of data supporting quantitative estimates, we assume that the waning rate of booster protection from infection is half the waning rate of the protection provided by second dose vaccine (see Figure 8B).

**Figure 8:**
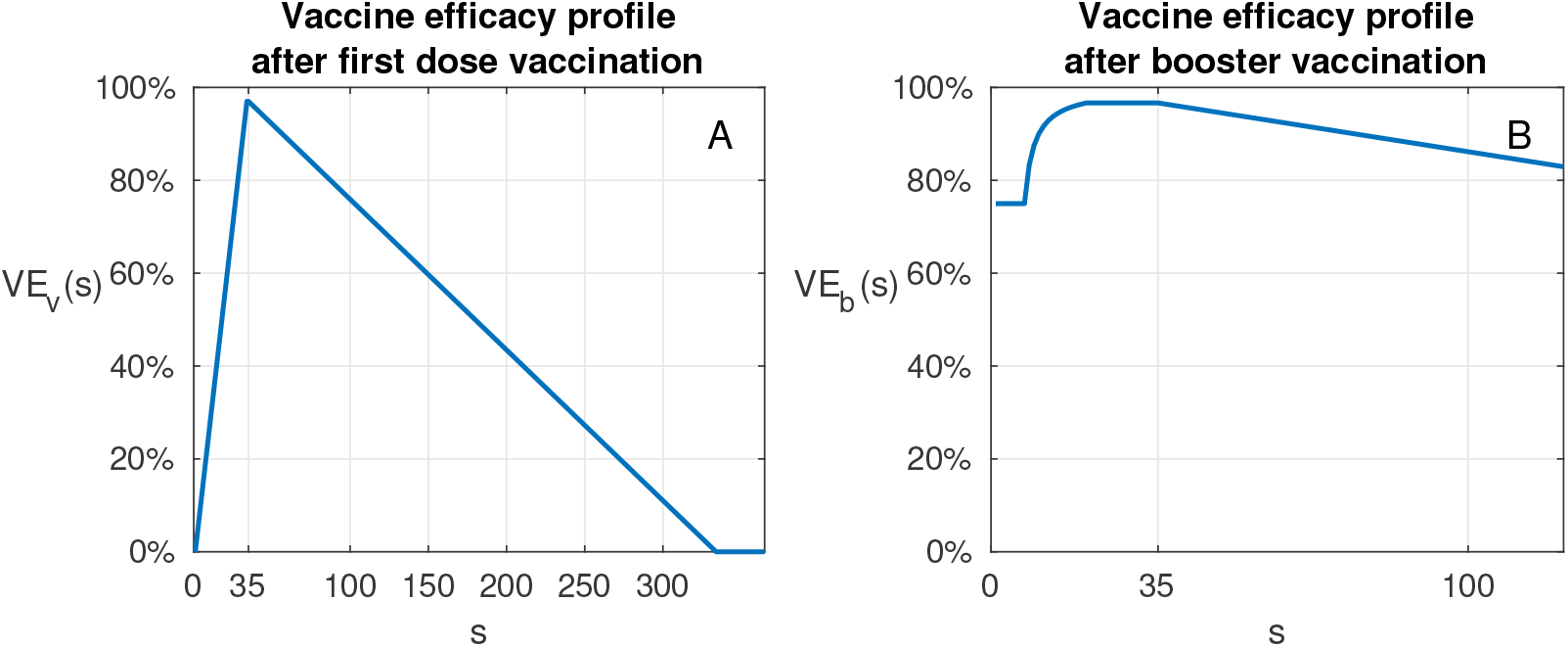
Vaccine efficacy profile from administration of first dose. A: Vaccine efficacy profile *VE*_*v*_(*s*) of individuals *s* days after receiving the first dose of vaccination. B: Vaccine efficacy profile *VE*_*b*_(*s*) of individuals *s* days after receiving booster vaccination.

The model assumes that the risk of a vaccinated person who has been infected to develop severe disease does not change in time. Thus the waning of vaccine efficacy against severe disease is due only to the waning of protection against infection. Under this assumption, the best fitted vaccine efficacy temporal profile well agrees with current estimates of vaccine efficacy against severe disease estimates [4], see Figure 7B.

##### Initialization

The initial values for the dynamic variables in the model were set as follows:

- *i*_*nv,j*_(*t*), (*t*_0_ *− τ*_*max*_ *≤ t ≤ t*_0_ *−* 1) - set as the non-vaccinated detected cases from age-group *j* with a 5-days lag (mean detection time) divided by the reporting rate *ρ*_*j*_ (see below).
- *i*_*v,j*_(*t*), (*t*_0_ *− τ*_*max*_ *≤ t ≤ t*_0_ *−* 1) - set as the vaccinated detected cases from age-group *j* with a 5-days lag divided by the reporting rate *ρ*_*j*_.
- *S*_*j*_(*t*_0_) - set as *N*_*j*_ minus the known non-vaccinated recovered cases up to time *t*_0_ divided by the reporting rate, minus the vaccinated up to time *t*_0_ and minus the infected at time *t*_0_.
- *V*_*j*_(*t*_0_) - set as the vaccinated from age-group *j* up to time *t*_0_ minus those infected after vaccination up to time *t*_0_.

Since the model was ran starting from July 1^st^ prior to the delivering of any booster shot, the variables *B* and *i*_*b*_ at time *t*_0_ were set to zero for all age-groups.

### C Model calibration

#### Observation of infections

The observation model connects the infection counts generated by the epidemic model to those observed in the data, taking into account the delay from infection to the identification of a case and the fact that only a fraction of infections are identified.

We denote by *ρ*_*j*_ the *reporting rates* for individuals of age group *j*, that is the probability that an infected case will be detected. We define the distribution of time from infection to detection by {*d*_*τ*_}, that is, *d*_*τ*_ is the probability that a person infected on day *t* will be detected on day *t* + *τ*, assuming detection.

The expected numbers of non-vaccinated, vaccinated, and boosted individuals detected in each age group on day *t* are then given by:

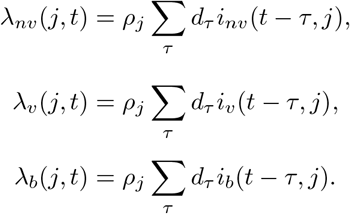

Denoting by 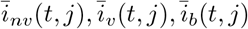, the numbers of infected individuals of age group *j* detected among the non-vaccinated, vaccinated, and booster-vaccinated classes on day *t*, we assume

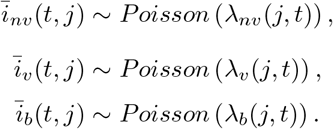

#### The likelihood function

Under this observation model, the likelihood corresponding to the infection data, given an epidemic trajectory generated by the epidemic model is:

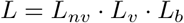

where

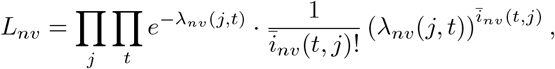

and similarly for *L*_*v*_, *L*_*b*_.

Since *L* depends on the epidemic model output for given parameter values, *L* is a function of these model parameters. The maximum likelihood estimates for the parameters are found by maximizing the function *L*, using numerical optimization. Overall, four parameters were estimated as part of fitting the data (Table 2). Detailed results of the model fit are shown in Figures 9-11. 95% CI for the parameter estimates were calculated using likelihood profiles [20] (Figure 15).

**Figure 9:**
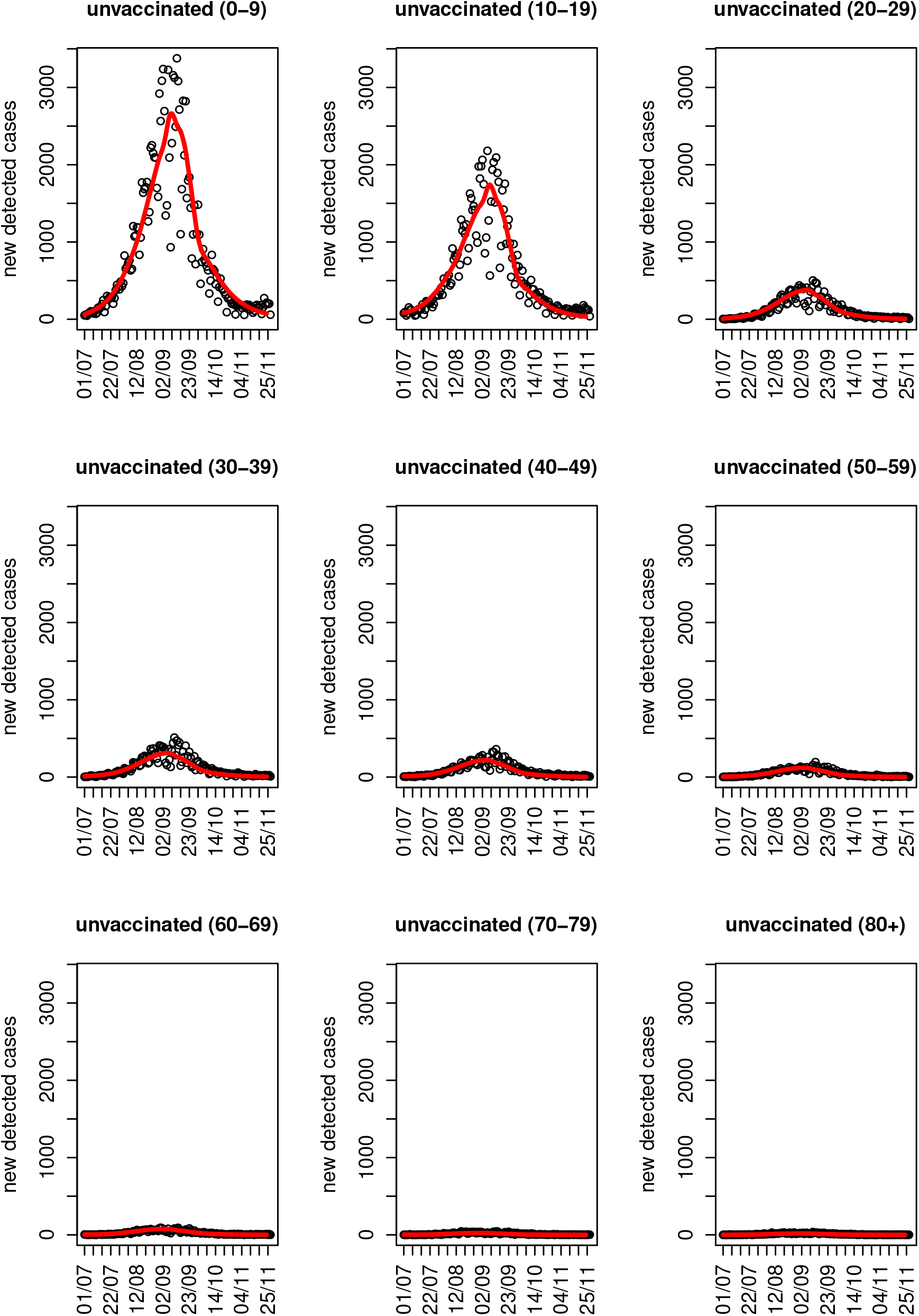
Model fit to detected cases incidences in unvaccinated individuals. Points indicate the observed data while the red lines indicate the model fit.

**Figure 10:**
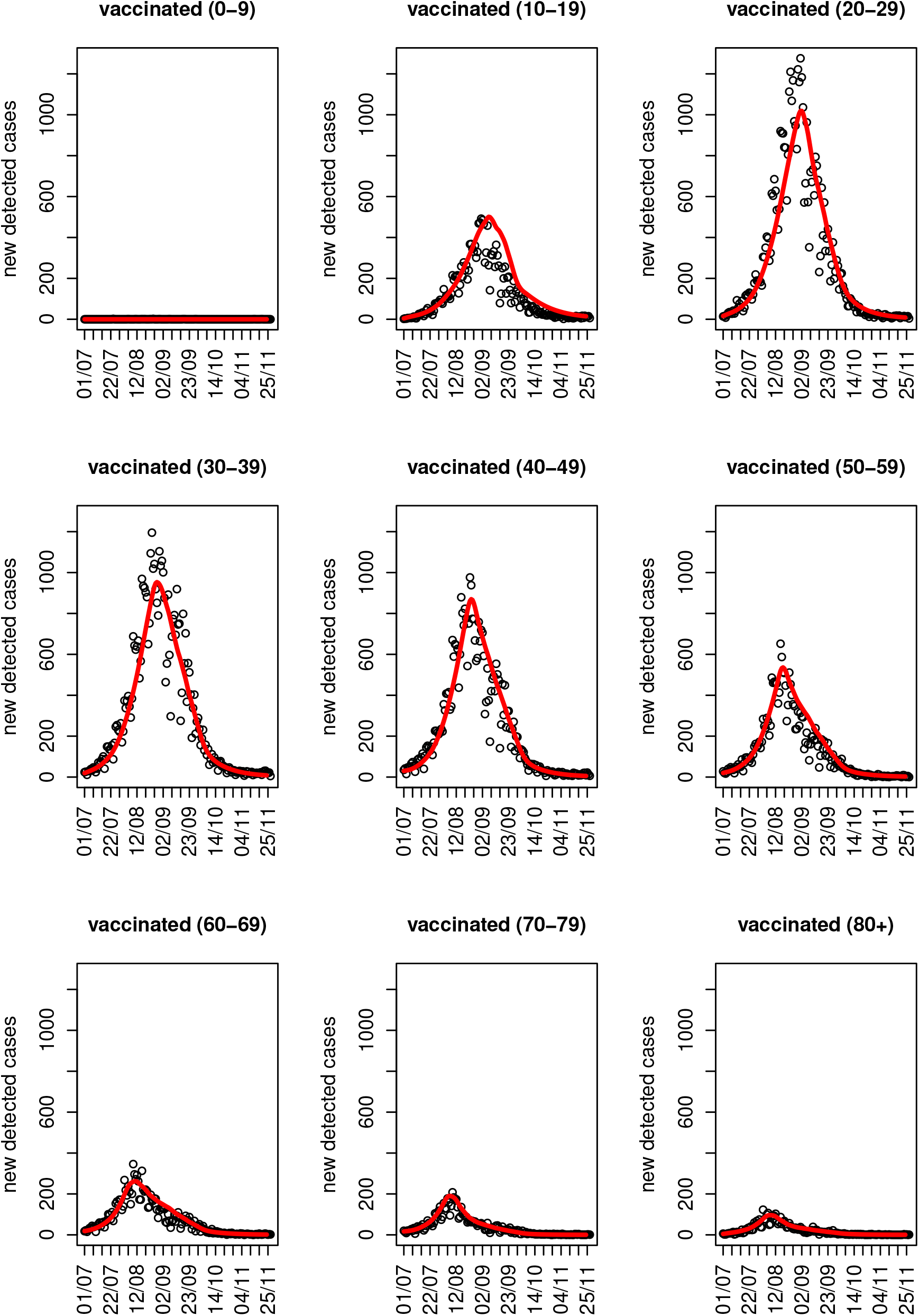
Model fit to detected cases incidences in vaccinated individuals. Points indicate the observed data while the red lines indicate the model fit.

**Figure 11:**
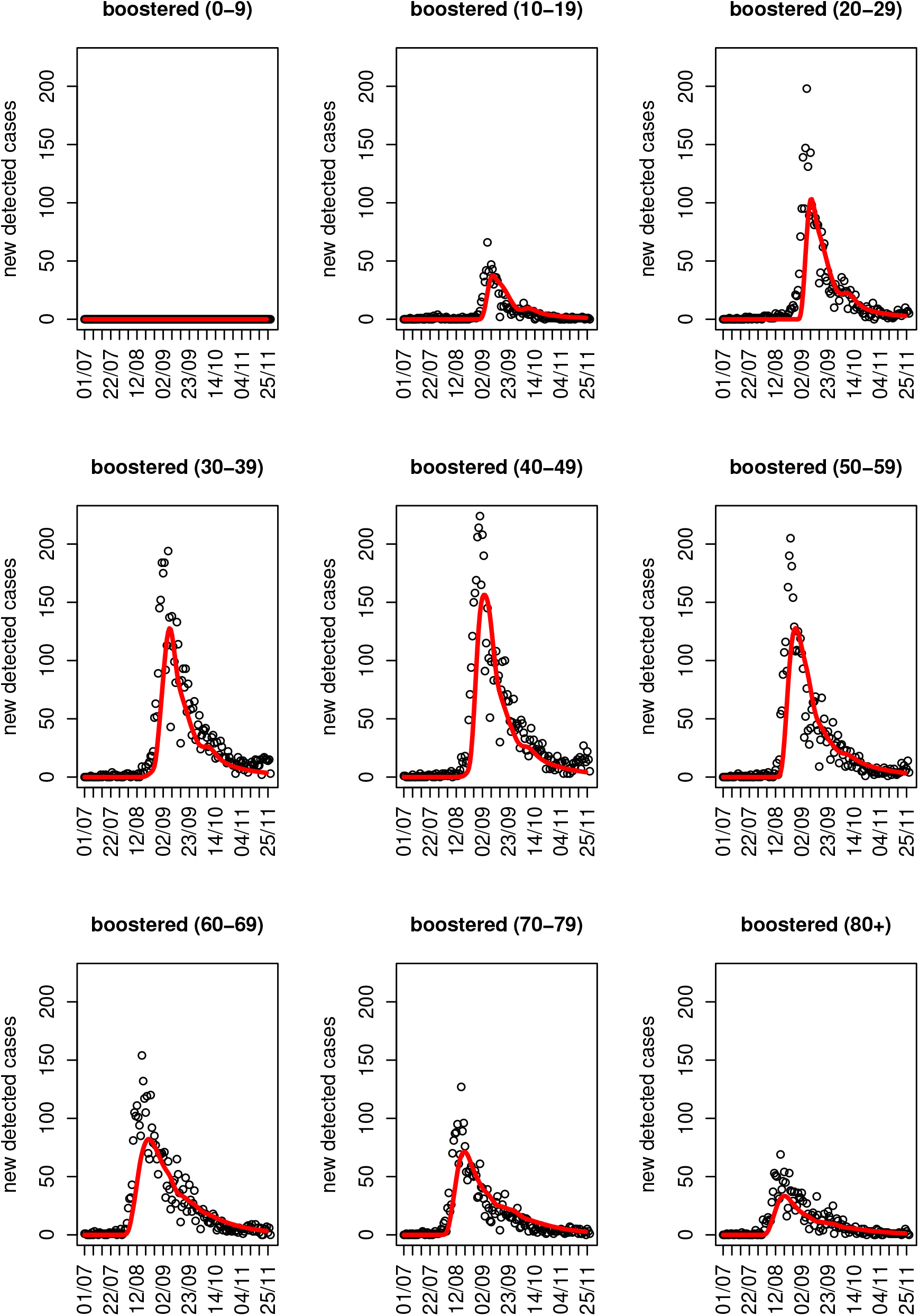
Model fit to detected cases incidences in boostered individuals. Points indicate the observed data while the red lines indicate the model fit.

**Figure 12:**
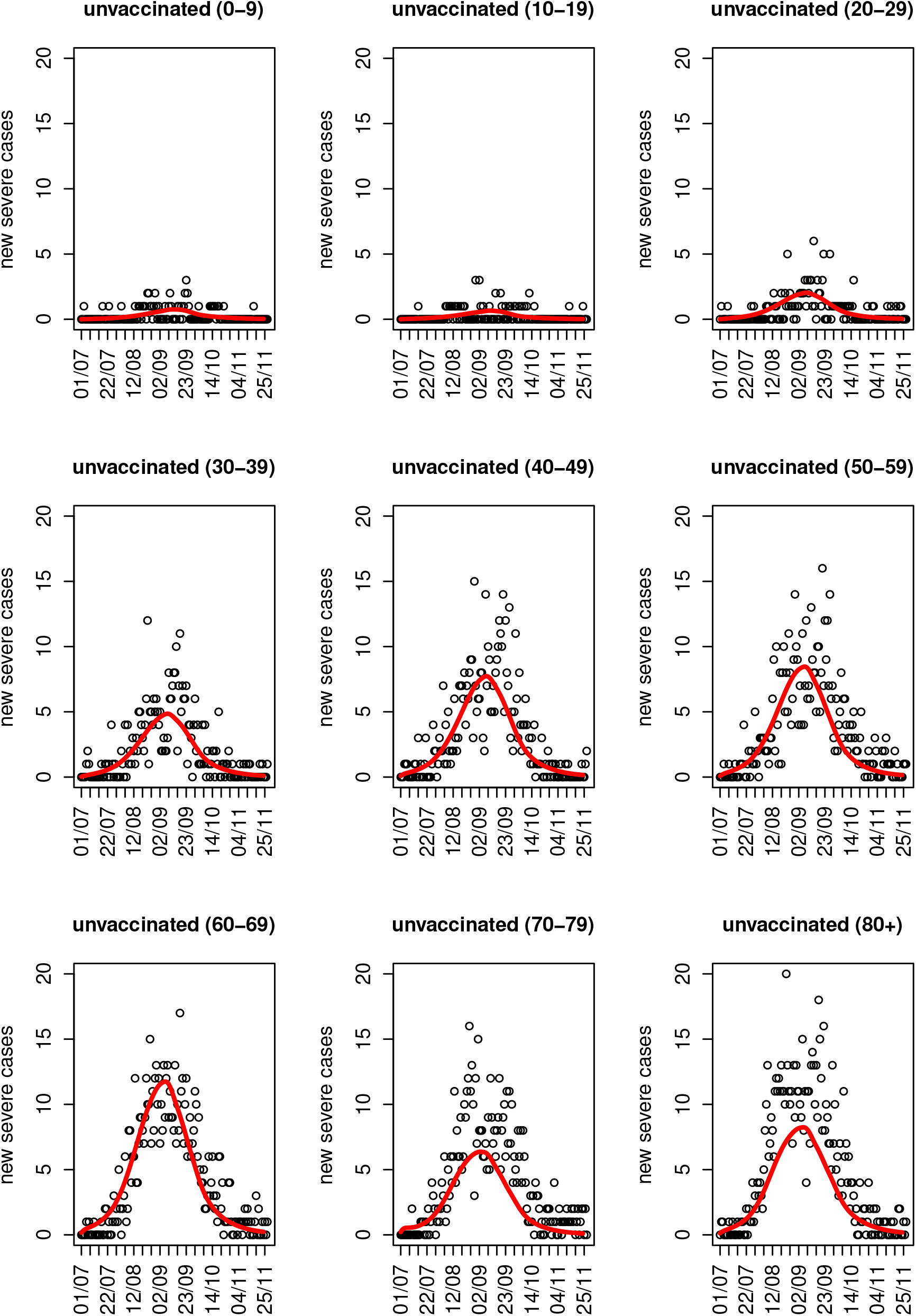
Model fit to severe cases incidences in unvaccinated individuals. Points indicate the observed data while the red lines indicate the model fit.

**Figure 13:**
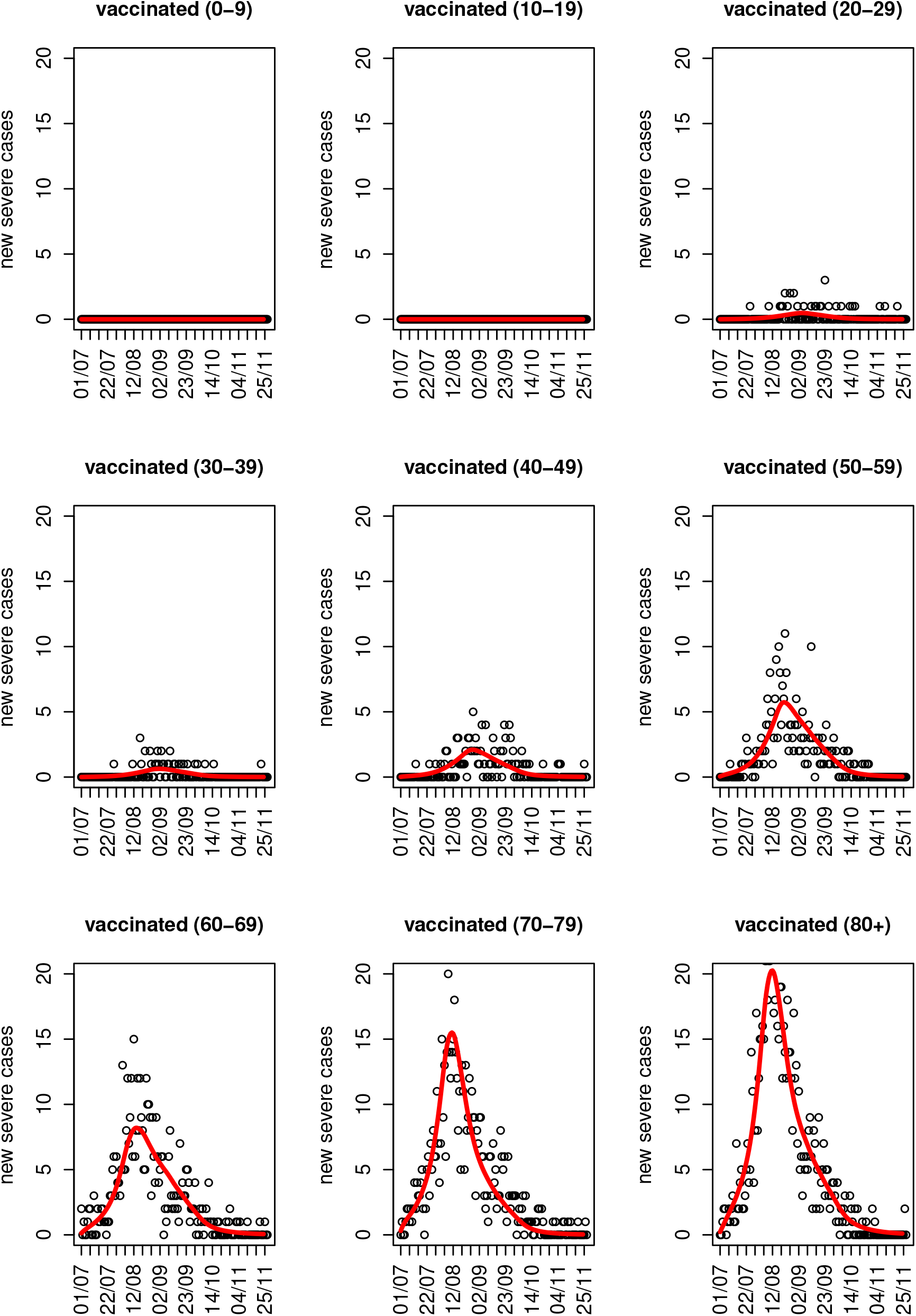
Model fit to severe cases incidences in vaccinated individuals. Points indicate the observed data while the red lines indicate the model fit.

**Figure 14:**
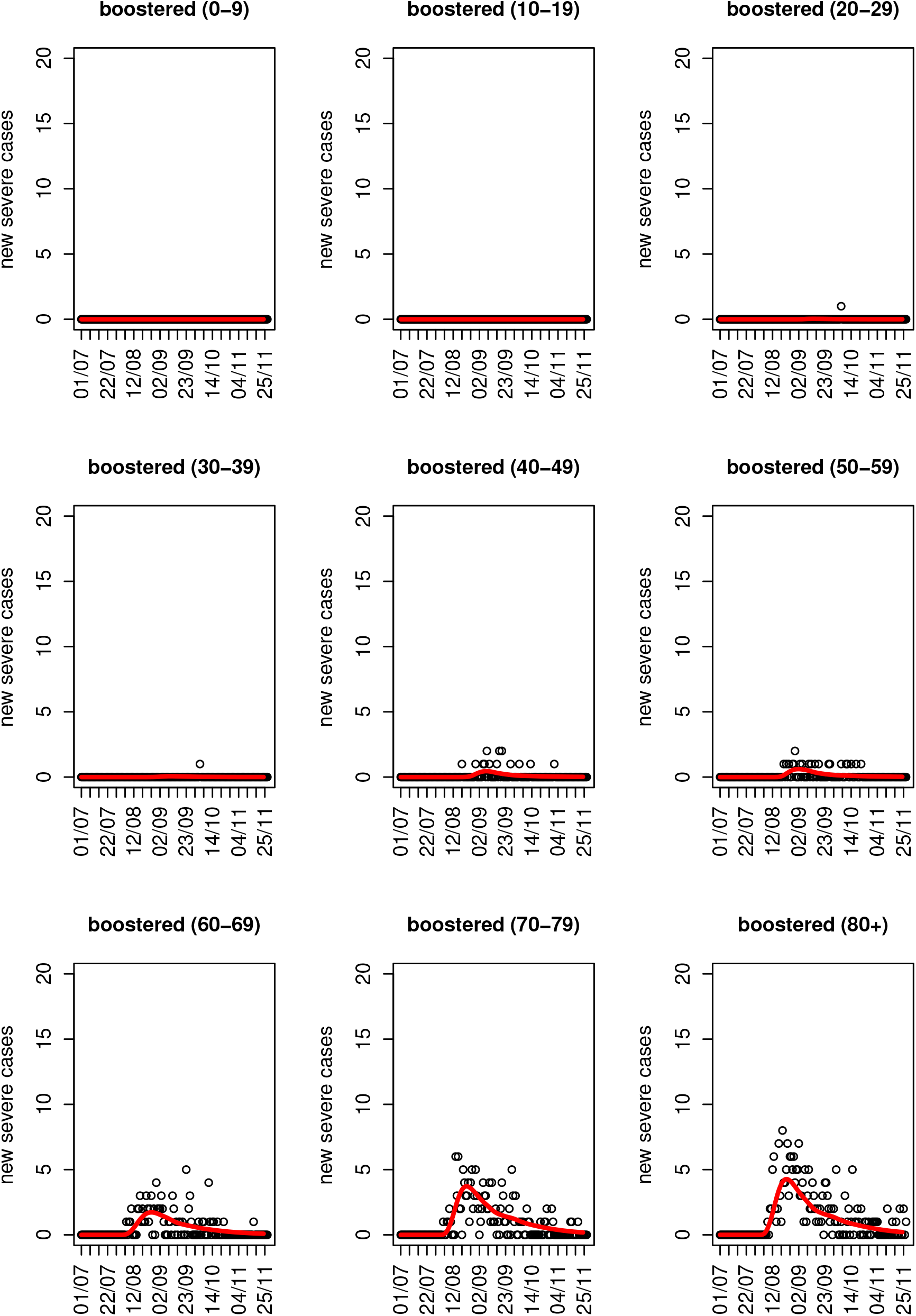
Model fit to severe cases incidences in boostered individuals. Points indicate the observed data while the red lines indicate the model fit.

**Figure 15:**
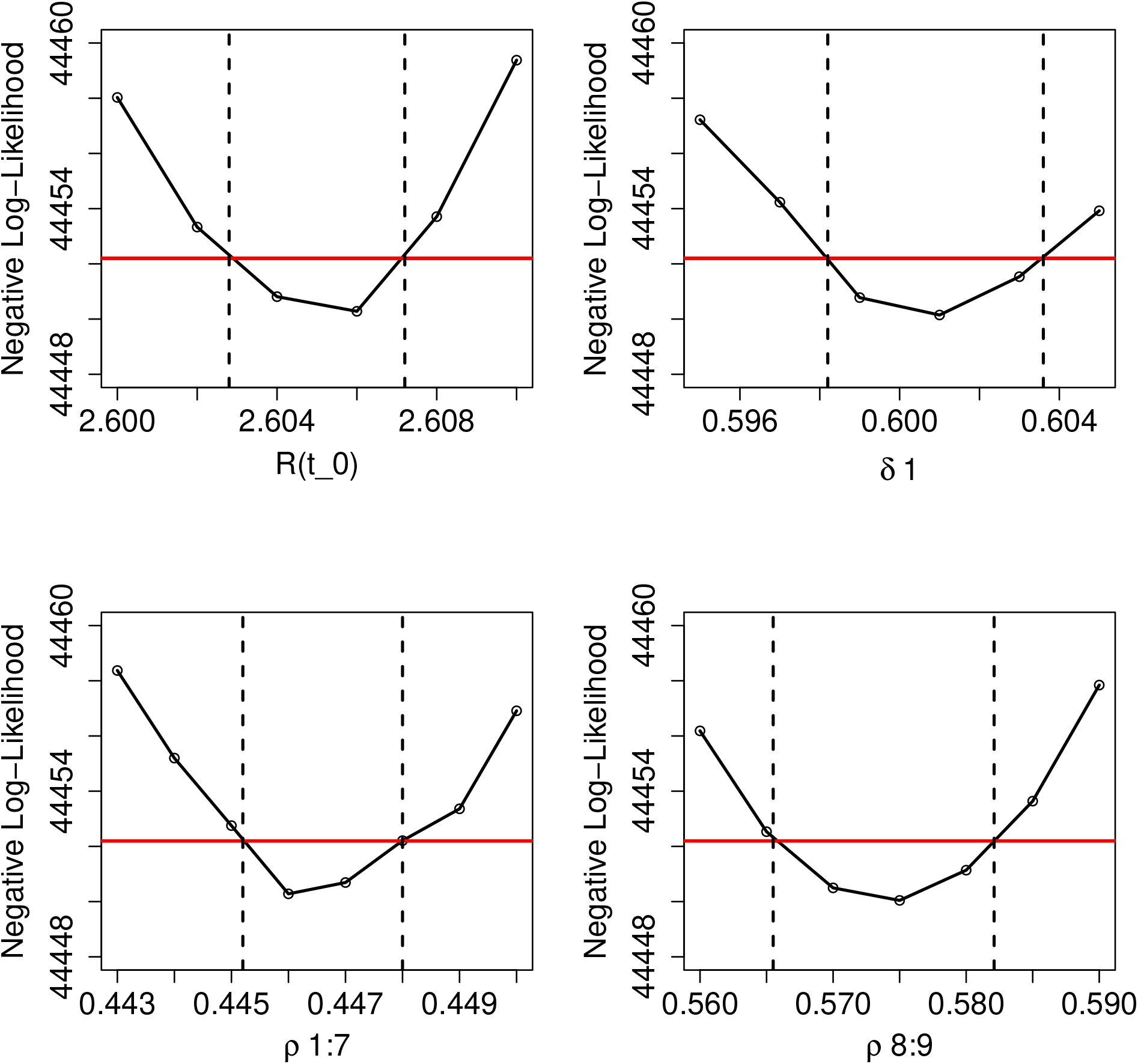
Likelihood profiles for the estimated parameters.

#### Modeling severe cases

We extend the model by modeling the flux of severe cases from detected cases. We denote by {*d*_*sev,τ*_} the distribution of time from infection to becoming severe, that is a person infected on day *t* who becomes a severe case, will become severe on day *t* + *τ* with probability *d*_*sev,τ*_. The probability that an unvaccinated detected person of age-group *j* will become a severe case is denoted by *κ*_*nv,j*_. Analogous probabilities for vaccinated individuals and individuals who have received a booster are denoted by *κ*_*v,j*_ and *κ*_*b,j*_. These probabilities were all extracted from the data (Table 2). We assume that all severe cases are detected.

The expected numbers of unvaccincated, vaccinated and booster-vaccinated from age-group *j* that reach a severe state at time *t* are then given by

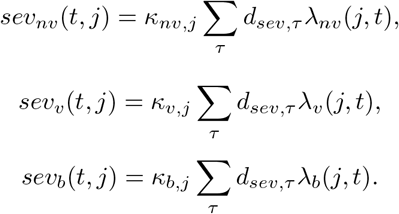

Setting the parameters to the maximum likelihood estimates obtained from fitting the detected cases, we run the extended model and compare the results to the observed incidences of new severe cases. Detailed results of the model fit to the these data are shown in Figures 12-14.

## References

[1] Noa Dagan, Noam Barda, Eldad Kepten, Oren Miron, Shay Perchik, Mark A Katz, Miguel A Hernán, Marc Lipsitch, Ben Reis, and Ran D Balicer. Bnt162b2 mrna covid-19 vaccine in a nationwide mass vaccination setting. New England Journal of Medicine, 2021.

[2] Eric J Haas, Frederick J Angulo, John M McLaughlin, Emilia Anis, Shepherd R Singer, Farid Khan, Nati Brooks, Meir Smaja, Gabriel Mircus, Kaijie Pan, et al. Nationwide vaccination campaign with bnt162b2 in israel demonstrates high vaccine effectiveness and marked declines in incidence of sars-cov-2 infections and covid-19 cases, hospitalizations, and deaths. 2021.

[3] Yair Goldberg, Micha Mandel, Yonatan Woodbridge, Ronen Fluss, Ilya Novikov, Rami Yaari, Arnona Ziv, Laurence Freedman, and Amit Huppert. Protection of previous sars-cov-2 infection is similar to that of bnt162b2 vaccine protection: A three-month nationwide experience from israel. medRxiv, 2021.

[4] Yair Goldberg, Micha Mandel, Yinon M Bar-On, Omri Bodenheimer, Laurence Freedman, Eric J Haas, Ron Milo, Sharon Alroy-Preis, Nachman Ash, and Amit Huppert. Waning immunity after the bnt162b2 vaccine in israel. New England Journal of Medicine, 2021.

[5] Yinon M Bar-On, Yair Goldberg, Micha Mandel, Omri Bodenheimer, Laurence Freedman, Nir Kalkstein, Barak Mizrahi, Sharon Alroy-Preis, Nachman Ash, Ron Milo, et al. Protection of bnt162b2 vaccine booster against covid-19 in israel. New England Journal of Medicine, 2021.

[6] COVID-19 Treatment Guidelines Panel. Coronavirus disease 2019 (covid-19) treatment guidelines. Accessed on December 2021.

[7] Sara Y Tartof, Jeff M Slezak, Heidi Fischer, Vennis Hong, Bradley K Ackerson, Omesh N Ranasinghe, Timothy B Frankland, Oluwaseye A Ogun, Joann M Zamparo, Sharon Gray, et al. Effectiveness of mrna bnt162b2 covid-19 vaccine up to 6 months in a large integrated health system in the usa: a retrospective cohort study. The Lancet, 398(10309):1407–1416, 2021.

[8] Dina Mistry, Maria Litvinova, Ana Pastore y Piontti, Matteo Chinazzi, Laura Fumanelli, Marcelo FC Gomes, Syed A Haque, Quan-Hui Liu, Kunpeng Mu, Xinyue Xiong, et al. Inferring high-resolution human mixing patterns for disease modeling. Nature communications, 12(1):1–12, 2021.

[9] Google. Covid-19 community mobility reports. www.google.com/covid19/mobility, 2021. [Online; accessed 20-December-2021].

[10] Hagai Rossman, Tomer Meir, Jonathan Somer, Smadar Shilo, Rom Gutman, Asaf Ben Arie, Eran Segal, Uri Shalit, and Malka Gorfine. Hospital load and increased covid-19 related mortality in israel. Nature Communications, 12(1):p1–7, 2021.

[11] Molly E Gallagher, Andrew J Sieben, Kristin N Nelson, Alicia NM Kraay, Walter A Orenstein, Ben Lopman, Andreas Handel, and Katia Koelle. Indirect benefits are a crucial consideration when evaluating sars-cov-2 vaccine candidates. Nature medicine, 27(1):4–5, 2021.

[12] Rachel Jordan, Martin Connock, Esther Albon, Anne Fry-Smith, Babatunde Olowokure, Jeremy Hawker, and Amanda Burls. Universal vaccination of children against influenza: are there indirect benefits to the community?: a systematic review of the evidence. Vaccine, 24(8):1047–1062, 2006.

[13] Martin Eichner, Markus Schwehm, Linda Eichner, and Laetitia Gerlier. Direct and indirect effects of influenza vaccination. BMC infectious diseases, 17(1):1–8, 2017.

[14] Yinon M. Bar-On, Yair Goldberg, Micha Mandel, Omri Bodenheimer, Laurence Freedman, Sharon Alroy-Preis, Nachman Ash, Amit Huppert, and Ron Milo. Protection against covid-19 by bnt162b2 booster across age groups. New England Journal of Medicine, 0(0):null, 0.

[15] Noam Barda, Noa Dagan, Cyrille Cohen, Miguel A Hernán, Marc Lipsitch, Isaac S Kohane, Ben Y Reis, and Ran D Balicer. Effectiveness of a third dose of the bnt162b2 mrna covid-19 vaccine for preventing severe outcomes in israel: an observational study. The Lancet, 2021.

[16] Ross J Harris, Jennifer A Hall, Asad Zaidi, Nick J Andrews, J Kevin Dunbar, and Gavin Dabrera. Effect of vaccination on household transmission of sars-cov-2 in england. New England Journal of Medicine, 2021.

[17] Kiesha Prem, Kevin van Zandvoort, Petra Klepac, Rosalind M Eggo, Nicholas G Davies, Centre for the Mathematical Modelling of Infectious Diseases COVID-19 Working Group, Alex R Cook, and Mark Jit. Projecting contact matrices in 177 geographical regions: an update and comparison with empirical data for the covid-19 era. PLoS computational biology, 17(7):e1009098, 2021.

[18] Yair Goldberg. personal communication.

[19] Fernando P Polack, Stephen J Thomas, Nicholas Kitchin, Judith Absalon, Alejandra Gurtman, Stephen Lockhart, John L Perez, Gonzalo Pérez Marc, Edson D Moreira, Cristiano Zerbini, et al. Safety and efficacy of the bnt162b2 mrna covid-19 vaccine. New England Journal of Medicine, 383(27):2603–2615, 2020.

[20] Benjamin M Bolker. Ecological models and data in R. Princeton University Press, 2008.

